# Multi-Omics characterization of biological pathways linking healthy dietary patterns to cardiometabolic disease risk across diverse populations

**DOI:** 10.64898/2026.02.23.26346874

**Authors:** Jihee Han, Keyong Deng, Zhen Hong, Zheqing Zhang, Nastya Godneva, Renée de Mutsert, Astrid van Hylckama Vlieg, Frits R. Rosendaal, Dennis O. Mook-Kanamori, Ju-Sheng Zheng, Yuming Chen, Eran Segal, Ruifang Li-Gao, DIYUFOOD consortium

## Abstract

**Background and Objectives:** Recent large-scale studies have consistently linked healthy dietary patterns to improved cardiometabolic health; however, the underlying biological pathways remain largely unclear, especially in non-European populations. In this study, we leverage data from four population-based cohorts (UK Biobank, NEO study, GNHS, and 10K) to investigate both common and cohort-specific biological pathways linking healthy dietary patterns to cardiometabolic disease through multi-omics profiling.

**Material and methods:** In each cohort, we first assessed the associations between each of the five major dietary pattern scores (i.e., AMED, hPDI, DII, AHEI, and EDIH) and cardiometabolic disease risk using Cox or logistic regression models. To explore the potential mediating role, metabolomics and proteomics measurements were incorporated into the models. All models were adjusted for relevant confounders, and false discovery rate correction was applied to account for multiple testing.

**Results:** With a total of 71,679 individuals without pre-existing cardiometabolic disease across four participating cohorts (UKB: 54,024, NEO: 4,838, GNHS: 3,201, and 10K: 9,616), we confirmed that adherence to healthy dietary patterns was associated with a 5-10% reduced risk of cardiometabolic disease. Three common biological pathways were identified: (1) mediation via large HDL particles and apolipoprotein F; (2) mediation via DNAJ/Hsp40 and triglyceride-rich lipoproteins; and (3) mediation via CRHBP-regulated HPA axis activity affecting triglyceride-rich lipoproteins.

**Conclusions:** Our integrative multi-omics analysis across diverse populations identifies novel biomarkers that connect healthy dietary patterns with cardiometabolic risk. These findings deepen our understanding of the biological mechanisms underlying diet-related disease and hold promise for enhancing the development of precision nutrition interventions.

## Introduction

A dietary pattern refers to the overall combination of foods and beverages that people habitually consume. Rather than focusing on individual nutrients or single food items, dietary patterns consider the synergistic effects of the entire diet. Based on cultural, geographical, and methodological factors, many dietary patterns were defined in the past decade, such as Mediterranean and plant-based dietary patterns. Recent large-scale observational studies focused on nine major dietary patterns and demonstrated that higher dietary pattern scores, indicating an overall healthier diet, are associated with lower risks of cardiometabolic diseases (i.e., cardiovascular disease and type 2 diabetes) and cancer^1,2^, as well as healthy aging^3^.

While these findings underscore the importance of overall diet quality, understanding how dietary patterns influence disease risk remains a critical next step. Identifying the biological pathways that mediate the relationship between diet and health outcomes, such as inflammation, insulin resistance, lipid metabolism, and gut microbiota, can provide valuable insights into the mechanisms underlying these associations. Advances in omics technologies have enabled profiling of proteomics and metabolomics alterations, which offers more comprehensive insight into the biological pathways linking a healthy diet to clinical outcomes. However, most studies have focused on a single omics layer, such as proteomics, in a single population. For example, Zhu et al. examined plasma proteomic signatures of healthy dietary patterns in relation to cardiometabolic diseases in UK populations only^4^, while Wang et al. explored metabolomic markers of Mediterranean diet adherence in European cohorts without proteomic integration^5^. These approaches limit our understanding of the complex interplay between diet and disease across diverse populations and multiple biological systems.

To address these gaps, our study uses data from four independent cohorts with diverse ethnicities, lifestyles, and socioeconomic statuses to investigate the biological pathways that mediate the associations between five major dietary patterns and cardiometabolic disease risk. We apply a multi-omics approach, i.e., integrating metabolomics and proteomics biomarkers, and perform Mendelian randomization analysis to uncover mechanistic insights into these diet-disease relationships. To assess the consistency of diet-disease associations across diverse genetic, cultural and lifestyle backgrounds, we analyzed data from four population-based cohort studies: the Netherlands Epidemiology of Obesity (NEO) study^12^, the UK Biobank (UKB)^13^, the 10K study^14^, and the Guangzhou Nutrition and Health Study (GNHS)^15^, that represent Dutch, British, Israeli, and Chinese populations, respectively.

We hypothesized that: 1) adherence to healthy dietary patterns is associated with a lower cardiometabolic disease risk across diverse populations; 2) specific metabolites and proteins are consistently associated with dietary patterns across cohorts; and 3) these biomarkers mediate the associations between specific dietary patterns and disease risk by acting through distinct biological pathways. To test these hypotheses, we focused on five dietary pattern scores, namely Alternate Mediterranean Diet (AMED)^6^, Healthful Plant-Based Diet Index (hPDI)^7,8^, Dietary Inflammation Index (DII)^9^, Empirical Dietary Index for Hyperinsulinemia (EDIH)^10^, and Alternate Healthy Eating Index (AHEI)^11^. By integrating diverse populations with multi-omics data, we provide a comprehensive overview of molecular mechanisms by which adherence to healthy diets influences disease risk.

## Results

### Analytical framework and characteristics of participating cohorts

Our study employed a two-stage analytical strategy to maximize the utility of the data. The primary analysis (stage 1), assessing diet-disease associations, included all four cohorts (prospective analysis in the UKB and NEO; cross-sectional analysis in GNHS and 10K). The mechanistic investigation (stage 2), involving proteomics and metabolomics, was necessarily restricted to participant subsets with available metabolomic and proteomic data. **Figure 1** provides a graphical overview of the analytical framework, and **Figure S1** presents a flow diagram of participant selection across the four cohorts.

**Figure 1.**
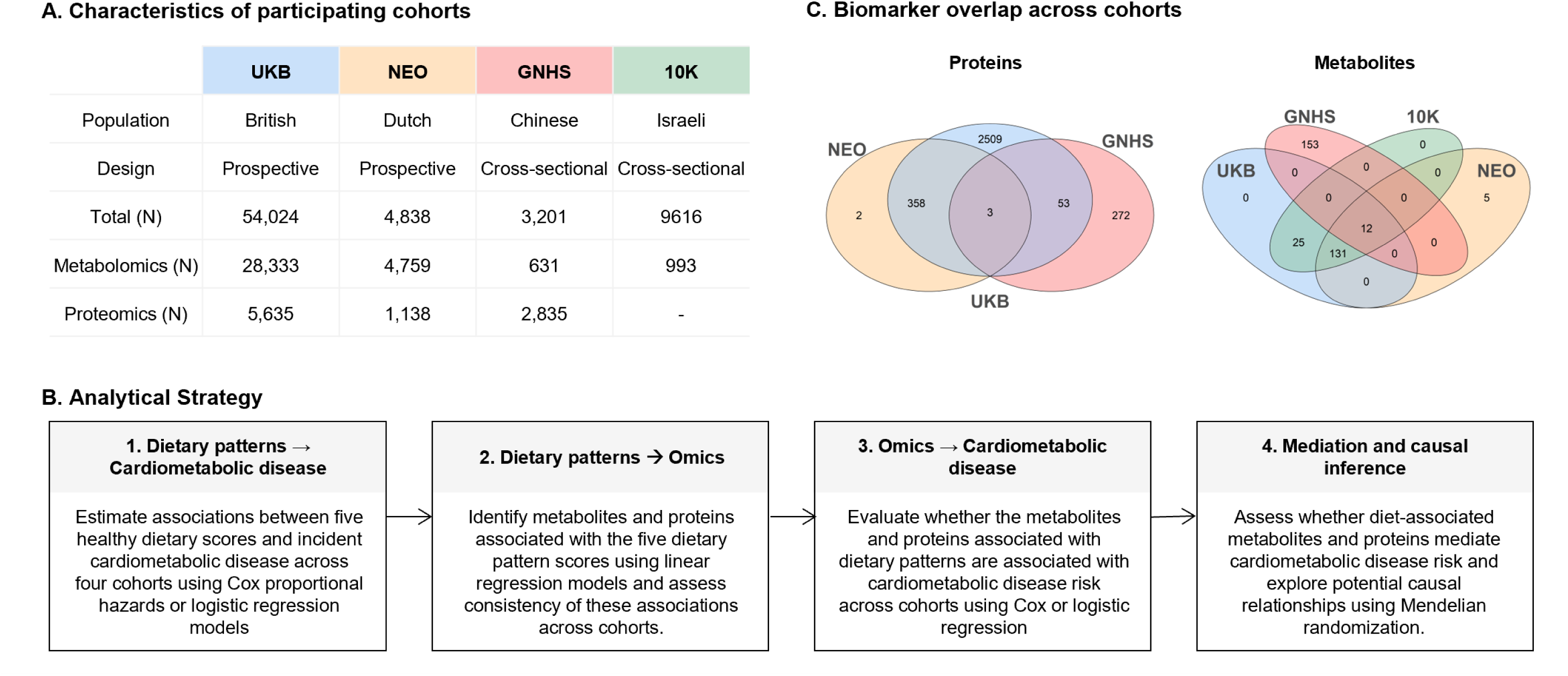
Overview of participating cohorts and analytical framework. (A) Overview of four population-based cohorts: the Netherlands Epidemiology of Obesity (NEO) study, the UK Biobank (UKB), the 10K study, and the Guangzhou Nutrition and Health Study (GNHS). (B) Analytical framework integrating epidemiological and multi-omics approaches. (C) Venn diagrams showing the overlap of proteins (left) and metabolites (right) associated with dietary patterns across cohorts.

A total of 71,679 individuals without pre-existing cardiometabolic diseases were included across different participating cohorts (UKB: 54,024, NEO: 4,838, GNHS: 3,201, and 10K: 9,616), with median ages ranging from 51 to 58 years (**Table 1**). Body mass index (BMI) varied considerably, with NEO showing the highest median (29.1kg/m²) due to oversampling of individuals with BMI >27kg/m², and GNHS the lowest (23.2kg/m²). These cohorts represent substantial ethnic and lifestyle diversity, such as physical activity levels (ranging from 11.6 MET-hours/week in 10K to 35.6 MET-hours/day in GNHS). While this difference is partly attributable to variations in the definitions of lifestyle factors across cohorts (see **Supplemental Material**), the observed population variations also reflect the diverse ethnic, cultural, and lifestyle backgrounds of the four cohorts, which enhances the generalizability of our findings.

**Table 1.** Baseline characteristics of study participants across four cohorts. Values are presented as mean (standard deviation), median (interquartile range), or number (percentage). Hormone replacement therapy users were counted exclusively among women. a. UKB use Townsend deprivation index for socioeconomic status, and the other cohorts use education levels. b. Only 170 of 3201 (5%) consumed alcohol (minimum = 0 and maximum 119.84). c. GNHS reported on per day and the other cohorts reported on per week. d. No vitamin use information was available in this subpopulation in UKB. NEO: Netherlands Epidemiology of Obesity study; GNHS: Guangzhou Nutrition and Health Study; 10K: Israel 10K Cohort; UKB: UK Biobank; AHEI: Alternate Healthy Eating Index; AMED: Alternate Mediterranean Diet; hPDI: Healthful Plant-Based Diet Index; rDII: reversed Dietary Inflammatory Index; rEDIH: reversed Empirical Dietary Index for Hyperinsulinemia; BMI: Body mass index.

| Characteristics | UKB | NEO | GHNS | 10K |
| --- | --- | --- | --- | --- |
| N | 54024 | 4838 | 3201 | 9616 |
| Age (years) | 57 (49,63) | 56 (50,61) | 58 (54,62) | 51 (46,57) |
| Sex (woman, %) | 29416 (54.4) | 2625 (54.3) | 2173 (67.9) | 5044 (52.5) |
| BMI (kg/m<sup>2</sup>) | 26.3 (23.7-29.3) | 29.1 (26.9, 2.0) | 23.2 (21.2, 25.2) | 25.6 (23.2, 28.5) |
| Smoking (current, %) | 4402 (8.1) | 766 (15.8) | 525 (16.4) | 1012 (10.5) |
| Socioeconomic status<sup>a</sup> |  |  |  |  |
| Education (>13 years, %) | - | 1877 (38.8) | 808 (25.2) | 6715 (69.8) |
| Townsend deprivation index | -1.4 (2.8) | - | - | - |
| Sleep duration (hours/day) | 7.0 (7.0, 8.0) | 7.0 (6.0, 7.8) | 8.0 (7.5, 9.0) | 7.0 (6.0,7.0) |
| Alcohol consumption (g/day) | 0.1 (0.0, 26.8) | 9.6 (2.1, 21.7) | 0.0 (0.0, 0.0)<sup>b</sup> | 0.6 (0.0,2.9) |
| Physical activity (MET-hours/week or day) | 31.1 (14.9, 60.2) | 27.5 (14.0, 47.3) | 35.6 (30.6, 47.6)<sup>c</sup> | 11.6 (4.4,23.1) |
| Vitamin use (yes, %)<sup>d</sup> | - | 1760 (36.4) | 701 (21.9) | 1788 (18.6) |
| Hormone replacement therapy (yes, %) | 2970 (10.1) | 69 (2.6) | 114 (3.6) | 981 (10.2) |
| Energy-adjusted score |  |  |  |  |
| AHEI | 43.0 (35.4, 50.7) | 42.1 (35.9, 48.8) | 49.7 (45.1, 54.3) | 52.0 (44.6,59.0) |
| AMED | 3.2 (2.2, 4.3) | 4.0 (2.9, 5.2) | 4.9 (3.8, 6.0) | 4.1 (2.9,5.3) |
| hPDI | 51.6 (48.3, 55.1) | 54.3 (50.2, 58.7) | 39.7 (35.2, 44.0) | 50.4 (46.2,54.9) |
| rDII | -0.6 (-1.7, 0.6) | 0.0 (-0.3, 0.4) | 3.2 (2.0, 4.1) | -1.3 (-1.9, -0.6) |
| rEDIH | -0.2 (-0.5, 0.1) | -0.6 (-1.1, -0.2) | 0.8 (0.5, 1.0) | -0.3 (-0.5, -0.2) |

### Consistent protective associations between healthy dietary patterns and cardiometabolic risk

Of the total population in each cohort, we observed 377 cardiometabolic disease cases newly diagnosed in the NEO study (median follow-up 6.6 years, interquartile range [IQR] = 5.8-7.4) and 4239 in UKB (median follow-up 12.7 years, IQR = 12.4-13.1) as well as 1497 and 1715 prevalent cases in GNHS and 10K, respectively. Overall, adherence to healthy dietary patterns was consistently associated with a lower risk of cardiometabolic disease across the four diverse populations, though the magnitude and precision of the associations varied (**Table 2**). After adjustment for confounders, all dietary pattern scores in UKB showed inverse associations (hazard ratios [HRs] ranging from 0.88 to 0.93 per standard deviation [SD]), with the strongest protective association observed for rEDIH (HR = 0.88, 95% CI = 0.86-0.91). Similarly, in the NEO study and 10K cohort, most dietary patterns showed protective associations, with rDII demonstrating the strongest association (HR = 0.87, 95% CI = 0.79-0.97; odds ratio [OR] = 0.89, 95% CI = 0.84-0.96). In GNHS, associations were generally neutral with ORs close to 1.00 (OR range = 0.97-1.00). After additional adjustment for BMI or family history of disease, the associations were generally attenuated but still showed the same directions.

**Table 2.** Associations between dietary pattern scores and cardiometabolic disease risk. Hazard ratios in UKB and NEO study or odds ratios in GNHS and 10K, with 95% confidence interval per standard deviation increase in dietary pattern scores across cohorts. Models were adjusted for age, sex, smoking, physical activity, education, vitamin use, hormone therapy, sleep duration, and alcohol (except for hPDI). AHEI: Alternate Healthy Eating Index; AMED: Alternate Mediterranean Diet; hPDI: Healthful Plant-Based Diet Index; rDII: reversed Dietary Inflammatory Index; rEDIH: reversed Empirical Dietary Index for Hyperinsulinemia; BMI: Body mass index; FH: Family history.

| Study | UKB | NEO | GNHS | 10K |
| --- | --- | --- | --- | --- |
| Person-years | 655289.4 | 30517.91 | - | - |
| Cases/total | 4239/54024 | 377/4838 | 1497/3201 | 1715/9616 |
| Effect estimates | HR (95% CI) | HR (95% CI) | OR (95% CI) | OR (95% CI) |
| AHEI |  |  |  |  |
| Crude | 0.90 (0.87-0.93) | 0.88 (0.79-0.97) | 0.97 (0.90, 1.04) | 0.98 (0.93, 1.04) |
| Adjusted | 0.89 (0.87-0.92) | 0.90 (0.81-1.00) | 0.97 (0.90, 1.04) | 0.91 (0.84, 0.97) |
| Adjusted+BMI | 0.97 (0.94-1.00) | 0.94 (0.84-1.04) | 0.97 (0.90, 1.04) | 0.94 (0.87, 1.01) |
| Adjusted+FH | 0.90 (0.87-0.93) | 0.90 (0.81-1.00) | - | 0.93 (0.85, 1.01) |
| AMED |  |  |  |  |
| Crude | 0.89 (0.86-0.91) | 0.90 (0.82-1.00) | 1.00 (0.93, 1.07) | 1.00 (0.95, 1.06) |
| Adjusted | 0.91 (0.88-0.93) | 0.93 (0.83-1.03) | 1.00 (0.94, 1.08) | 0.90 (0.84, 0.97) |
| Adjusted+BMI | 0.96 (0.93-0.99) | 0.96 (0.87-1.07) | 1.00 (0.93, 1.07) | 0.93 (0.87, 1.00) |
| Adjusted+FH | 0.91 (0.88-0.94) | 0.93 (0.83-1.03) |  | 0.94 (0.86, 1.02) |
| hPDI |  |  |  |  |
| Crude | 0.92 (0.90-0.95) | 0.89 (0.80-0.99) | 0.97 (0.91, 1.04) | 1.01 (0.96, 1.07) |
| Adjusted | 0.93 (0.90-0.95) | 0.95 (0.85-1.06) | 0.99 (0.92, 1.07) | 0.92 (0.86, 0.99) |
| Adjusted+BMI | 0.98 (0.95-1.02) | 0.98 (0.88-1.10) | 0.98 (0.91, 1.05) | 0.96 (0.89, 1.03) |
| Adjusted+FH | 0.93 (0.90-0.96) | 0.95 (0.85-1.06) | - | 0.93 (0.86, 1.01) |
| rDII |  |  |  |  |
| Crude | 0.99 (0.96-1.02) | 0.89 (0.81-0.99) | 0.96 (0.90, 1.03) | 1.03 (0.98, 1.09) |
| Adjusted | 0.92 (0.89-0.95) | 0.87 (0.79-0.97) | 0.97 (0.90, 1.04) | 0.89 (0.83, 0.95) |
| Adjusted+BMI | 0.95 (0.92-0.98) | 0.90 (0.81-1.00) | 0.97 (0.90, 1.04) | 0.91 (0.85, 0.98) |
| Adjusted+FH | 0.92 (0.89-0.95) | 0.87 (0.79-0.97) | - | 0.91 (0.83, 0.99) |
| rEDIH |  |  |  |  |
| Crude | 0.87 (0.84-0.89) | 0.87 (0.79-0.95) | 1.00 (0.93, 1.07) | 0.94 (0.89, 0.99) |
| Adjusted | 0.88 (0.86-0.91) | 0.93 (0.85-1.03) | 1.00 (0.93, 1.07) | 0.95 (0.88, 1.02) |
| Adjusted+BMI | 0.93 (0.90-0.96) | 0.98 (0.88-1.08) | 1.00 (0.93, 1.08) | 0.98 (0.92, 1.06) |
| Adjusted+FH | 0.89 (0.86-0.91) | 0.93 (0.84-1.03) | - | 0.92 (0.84, 1.01) |

For type 2 diabetes (**Table S1**), UKB, NEO, and 10K consistently showed inverse associations across all dietary pattern scores. In UKB, rEDIH demonstrated the strongest protective association (HR = 0.84, 95% CI = 0.81-0.87), while AHEI showed the strongest associations in NEO, GNHS, and 10K. Associations for cardiovascular disease or dyslipidemia were consistently protective across all patterns in UKB and 10K, but less consistent in NEO and GNHS, with most confidence intervals including 1. For the subsequent mediation analysis, we further limited the study population to individuals with available proteomics (**Table S2)** and metabolomics data (**Table S3)**. This restriction resulted in attenuated effect sizes and wider confidence intervals for diet-disease associations in the omics subsets.

### Identification and cross-cohort replication of dietary pattern-associated multi-omic signatures

To identify the molecular mediators, we profiled proteomics and metabolomics. We focused on biomarkers that showed significant associations with at least one dietary pattern in the UKB (q < 0.05) and were replicated in at least one additional cohort (p < 0.05) with consistent effect directions. **Figure 2B** presents the number of proteins and metabolites overlapped across cohorts.

**Figure 2.**
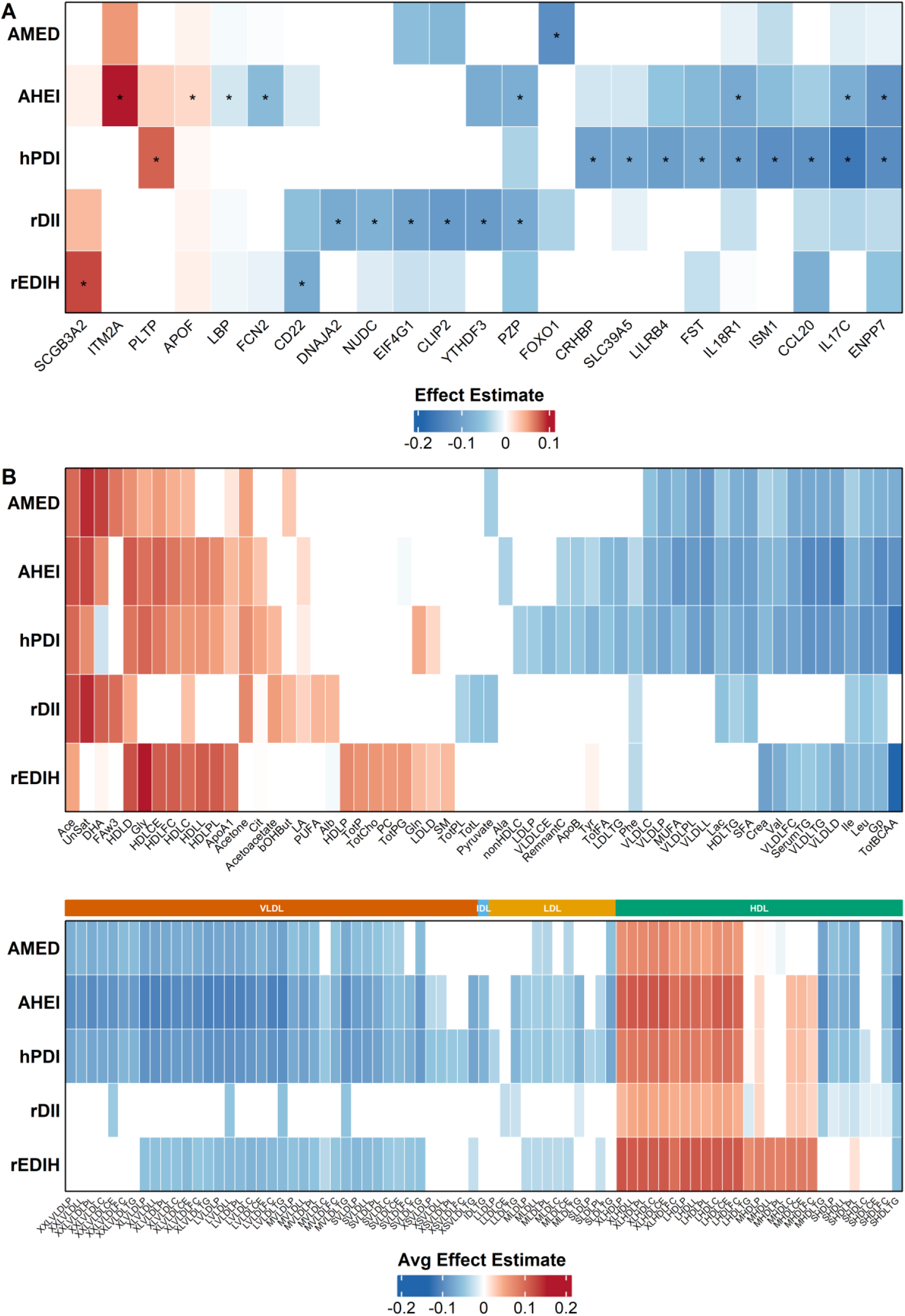
Cross-cohort replicated multi-omic signatures associated with five healthy dietary patterns. (A) Heatmap showing diet and protein associations in the NEO and GNHS cohorts, which shows consistent effect directions with the significant associations identified in the UKB (q < 0.05). Asterisks indicate associations with p < 0.05 in the replication cohorts. Color represents beta coefficients. (B) Heatmap showing the average effect estimates of significant diet–metabolite associations observed in at least two populations. The upper panel presents metabolites commonly measured across platforms, while the Lower panel focuses on lipoprotein subclasses. Colors represent standardized beta coefficients.

#### Proteomic signatures

Among the 414 eligible proteins, 244 were associated with at least one dietary pattern score in UKB, of which 23 proteins were replicated in at least one additional cohort (**Figure 2 and Table S4**). Replicated proteins primarily involve pathways related to transcription, cell development, inflammation, lipid metabolism, and protein structure (**Table S5**).

Several key proteins showed particularly consistent associations across dietary patterns. For example, ENPP7 (Ectonucleotide pyrophosphatase/phosphodiesterase family member 7), involved in fatty acid absorption in the intestine, was inversely associated with adherence to healthy dietary patterns in UKB and NEO cohorts. Similarly, APOF (Apolipoprotein F), involved in lipid metabolism, showed positive associations with adherence to healthy dietary patterns in UKB and GNHS cohorts, which suggests changes in the regulation of lipid transport and metabolism. These protein signatures provide mechanistic insights into how dietary patterns may influence biological pathways related to cardiometabolic health.

#### Metabolomic signatures

Among 168 eligible metabolites, 167 were associated with at least one dietary pattern score in UKB, with 146 replicating in at least one additional cohort (**Figure 2 and Table S6**). Several metabolites showed significant associations with all five dietary patterns across multiple cohorts, including acetate, leucine, isoleucine, lactate, and high-density lipoprotein (HDL) particles.

We observed consistent inverse associations between adherence to healthy dietary patterns and triglyceride levels, very-low-density lipoprotein (VLDL), and low-density lipoprotein (LDL) particles. Conversely, acetate, unsaturated fat, and large HDL particles demonstrated positive associations with adherence to healthy dietary patterns. These metabolomic signatures reflect biological pathways known to influence cardiometabolic health, including lipid metabolism, amino acid metabolism, and inflammation.

### Proteomic and metabolomic pathways mediating the protective effect

We performed mediation analysis to estimate the proportion of the association between each dietary pattern and cardiometabolic disease that was mediated through proteins and metabolites. We included proteins and metabolites associated with both dietary patterns and cardiometabolic disease and replicated across cohorts (**Table S7-S11 and Figure S2**).

#### Mediation through proteins

Across six proteins with mediating potential, CD22 mediated part of the association between the rEDIH and lower risks of cardiometabolic disease and type 2 diabetes, accounting for approximately 4–6% of the total effect in UKB. In addition, LILRB4 mediated the association between the hPDI and cardiometabolic disease and cardiovascular disease, accounting for around 20% of the total effect, although the confidence interval for cardiometabolic disease was wide, indicating substantial uncertainty. This protein is involved in immune regulation and inflammatory signaling, suggesting that the anti-inflammatory dietary pattern may improve cardiometabolic health partly through modulation of immune-related pathways. Other proteins, including APOF, CRHBP, EIF4G1, and DNAJA2, showed suggestive mediation effects (ranging from 7–40%) but with wide confidence intervals, indicating limited statistical precision and potential overestimation.

#### Mediation through metabolites

Among 90 replicated metabolites, 29 principal variables were selected using the iPVs approach to reduce redundancy among highly correlated metabolites and to capture the main metabolic dimensions representing overall metabolic variation (**Table S11**). Metabolites collectively accounted for substantial mediation proportions, explaining up to 37–61% of the protective effects of healthy dietary patterns on cardiometabolic disease. This finding suggests that metabolic changes in lipid metabolism represent important biological pathways through which dietary patterns influence cardiometabolic disease. Similar patterns were observed in NEO, although the confidence intervals were wide.

### Multi-omics integration through Mendelian randomization analyses

To investigate potential causal associations between metabolites and proteins in the dietary-cardiometabolic disease pathway, we conducted bidirectional Mendelian randomization analyses. After applying Bonferroni correction (**Figure 3**), metabolites such as large HDL and VLDL particles were significantly associated with higher levels of proteins (APOF, CRHBP, DNAJA2, and EIF4G1) using the inverse variance weighted (IVW) method. For instance, LHDLP was positively associated with APOF levels (β = 0.26, 95% CI = 0.16-0.37), and the association was supported by consistent results from sensitivity analysis (**Table S12)**. In the reverse analyses (protein as exposure and metabolite as outcome), none of the associations reached the Bonferroni-corrected threshold. However, effect estimates were similar to those of metabolites to proteins, even larger for APOF. This suggests a complex interplay that requires further investigation. Collectively, the Mendelian randomization findings support a cascade where adherence to healthy dietary patterns primarily alters lipid and amino acid metabolites, which subsequently influence key proteins.

**Figure 3.**
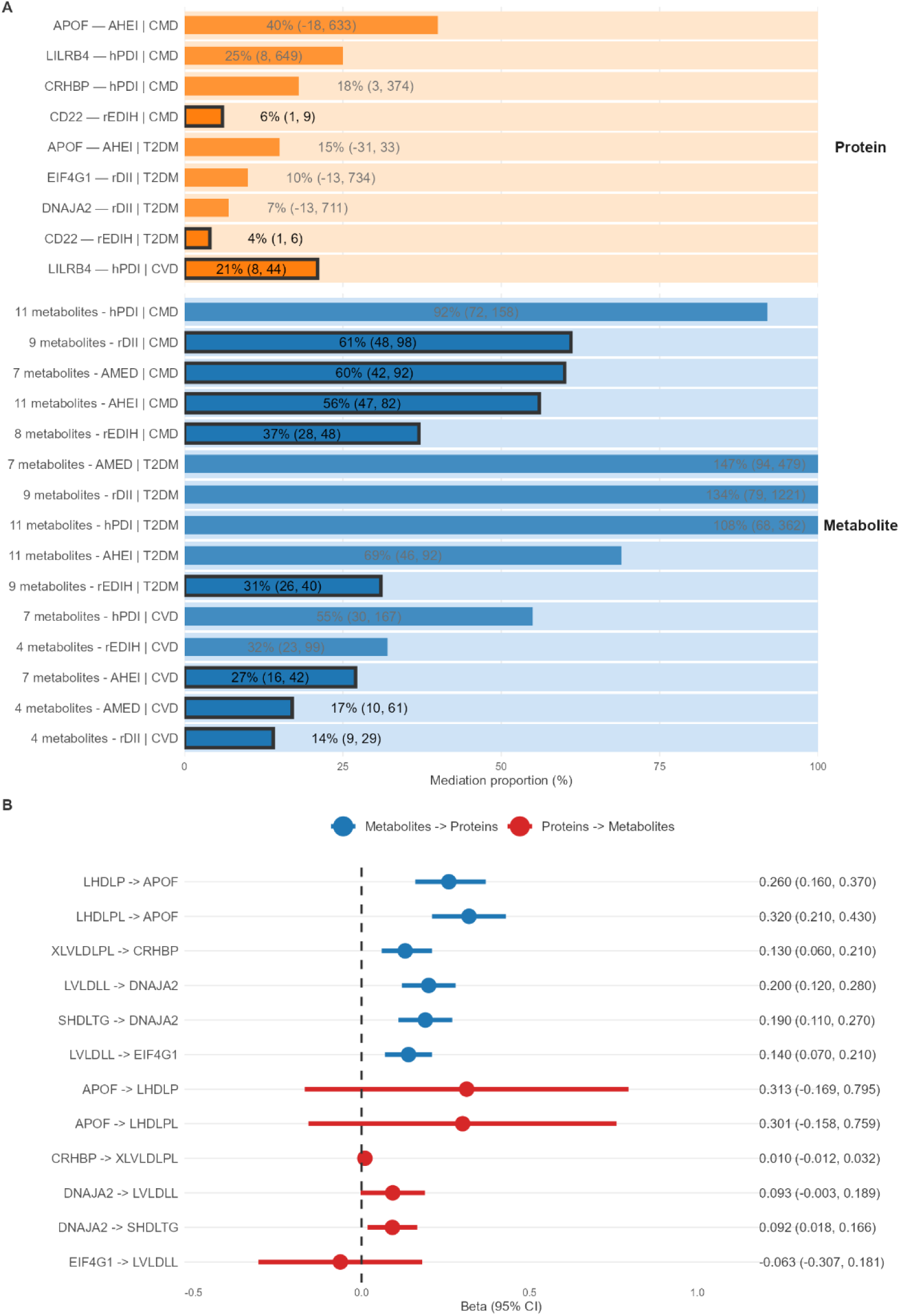
Mediation and causal relationships linking dietary patterns, circulating biomarkers, and cardiometabolic disease. (A) Mediation proportions (95% CI) for proteins and metabolites significantly associated with both dietary patterns and cardiometabolic disease outcomes in UKB. Key metabolites were selected using principal variable analysis. Cox proportional hazard models were adjusted for age, sex, smoking, physical activity, education, vitamin use, hormone therapy, sleep duration, and alcohol (except for hPDI). Mediation proportions represent the proportion of the total dietary effect on disease risk explained by mediators. (B) Bidirectional Mendelian randomization estimates for metabolite–protein associations passing Bonferroni correction (p < 5.56 × 10⁻⁴). Causal effects are shown as β coefficients (95% CI) from inverse-variance weighted analyses, with both forward (metabolite → protein) and reverse (protein → metabolite) directions presented. AHEI: Alternate Healthy Eating Index; AMED: Alternate Mediterranean Diet; hPDI: Healthful Plant-Based Diet Index; rDII: reversed Dietary Inflammatory Index; rEDIH: reversed Empirical Dietary Index for Hyperinsulinemia; CMD: cardiometabolic disease; T2DM: type 2 diabetes; CVD: cardiovascular disease.

### Biological pathways linking healthy dietary patterns to cardiometabolic disease

Integrating proteomic, metabolomic, and Mendelian randomization analyses provides insight into the molecular pathways through which dietary patterns may influence cardiometabolic health. As summarized in **Figure 4**, we identified three potential biological pathways linking healthy diets to a reduced risk of cardiometabolic disease. First, a common pathway involves interactions between large HDL particles (LHDL) and apolipoprotein F (APOF), suggesting a role for HDL-related lipid transport in mediating dietary effects on cardiometabolic disease risk. Second, a shared pathway links healthy dietary patterns to lower cardiometabolic disease risk through interactions between DNAJA2/Hsp40/EIF4G1 and triglyceride-rich lipoproteins, implicating protein homeostasis and lipid metabolism. Third, a distinct pathway connects healthy diets to reduced cardiometabolic disease risk via CRHBP-related modulation of the hypothalamic–pituitary–adrenal (HPA) axis, with downstream effects on triglyceride-rich lipoproteins.

**Figure 4.**
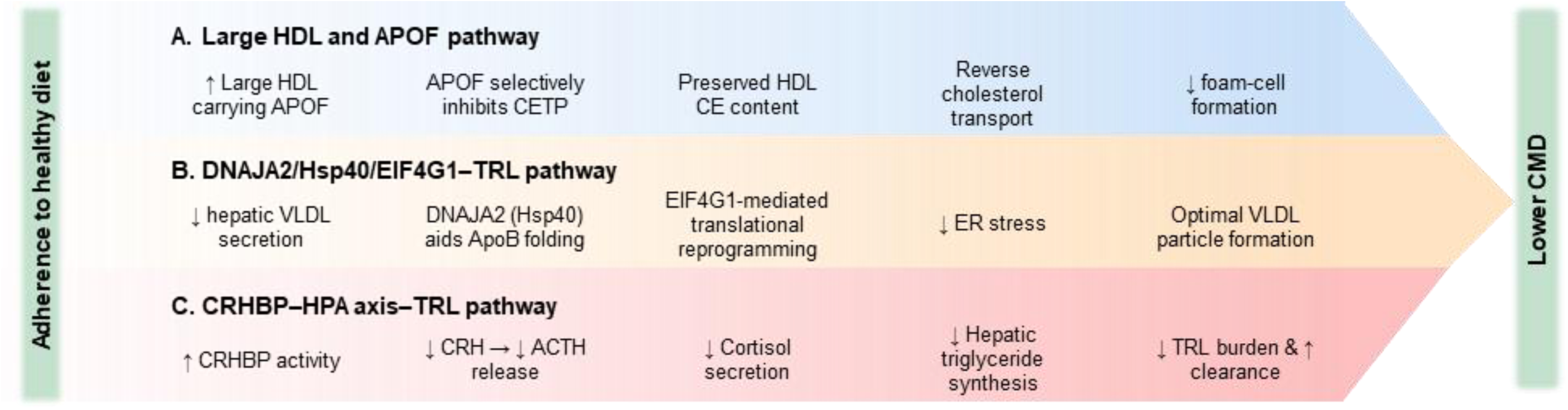
Integrated biological mechanisms linking healthy dietary patterns to reduced cardiometabolic disease risk. Schematic overview illustrating three convergent molecular pathways through which adherence to healthy dietary patterns may lower cardiometabolic disease (CMD) risk. (A) Large HDL and APOF pathway: Healthy dietary patterns increase the levels of large HDL particles (LHDL), which associate with apolipoprotein F (APOF). APOF-enriched HDL interacts with cholesterol-efflux receptors (ABCA1/ABCG1, SR-BI) and inhibits cholesteryl ester transfer protein (CETP) activity, preserving HDL cholesteryl ester (CE) content. This process enhances reverse cholesterol transport, reduces macrophage foam-cell formation and atherosclerosis, and ultimately decreases CMD risk. (B) DNAJA2/Hsp40–EIF4G1–TRL pathway: Healthy diets affect hepatic lipid metabolism, improving VLDL secretion balance and maintaining endoplasmic reticulum (ER) proteostasis. DNAJA2 (Hsp40) supports ApoB folding during VLDL assembly, while EIF4G1-mediated translational reprogramming (via the eIF2α–ATF4 axis) coordinates protein synthesis with chaperone activity. Together, these effects attenuate ER stress and inflammation, reducing CMD susceptibility. (C) CRHBP–HPA axis–TRL pathway: Healthy diets enhance corticotropin-releasing hormone–binding protein (CRHBP) activity, decreasing free CRH and subsequent ACTH-driven cortisol release. Reduced cortisol levels lower hepatic triglyceride synthesis and VLDL–ApoB100 assembly, leading to decreased TRL burden, improved lipid clearance, and diminished metabolic stress.

## Discussion

In this multi-cohort, multi-omics study of 71,679 participants from four diverse populations, we demonstrate that adherence to healthy dietary patterns is consistently associated with lower risk of cardiometabolic disease and identify robust molecular signatures that help explain these associations. Integrating plasma metabolomics and proteomics, we replicated a set of metabolite and protein markers across cohorts and found that metabolites account for a large portion of the protective effect of healthy diets, whereas individual proteins showed smaller but biologically plausible mediation. Mendelian randomization analyses further support a directional cascade in which diet-associated metabolites influence protein. Together, these findings provide mechanistic evidence linking dietary patterns to disease risk and highlight candidate metabolic and proteomic targets for further investigation.

### Confirmation of associations between healthy dietary patterns and lower cardiometabolic disease risks across diverse populations

Our findings regarding dietary patterns and cardiometabolic outcomes align with previous studies but extend them in important ways. The consistent inverse associations between adherence to healthy dietary patterns and cardiometabolic disease risk across populations with different genetic backgrounds, cultural contexts, and lifestyle factors strengthen the evidence for the beneficial effects of these dietary patterns beyond specific populations. Previous studies have predominantly focused on Western populations^1,2^, limiting their generalizability. Our inclusion of East Asian and Israeli populations addresses this limitation and suggests that the biological mechanisms linking diet to cardiometabolic health may be conserved across diverse populations. Nevertheless, we observed some variation in the strength of associations between dietary patterns and cardiometabolic outcomes across cohorts. These discrepancies may be attributable to both biological and methodological factors. Biologically, for instance, differences in genetic background or gut microbiome composition could modify diet and disease associations across populations. Methodologically, differences in dietary assessment tools, outcome definitions (incident disease versus phenotypes at baseline), follow-up duration, and sample sizes likely contributed to these variations. Additionally, cultural differences in food preparation methods and nutrient content of similarly named foods may have introduced unmeasured heterogeneity in dietary exposure classification despite our standardization efforts.

### Biological pathways linking healthy dietary patterns to cardiometabolic disease

The consistent inverse associations between adherence to healthy dietary patterns and inflammatory markers (e.g., glycoprotein acetyls), atherogenic lipid particles (e.g., VLDL, small LDL), and branched chain amino acid align with previous research linking these biomarkers to cardiometabolic disease^31^. Similarly, the positive associations between adherence to healthy dietary patterns and HDL particles, particularly large HDL, are consistent with the protective effects of these lipoproteins on cardiovascular risk^32^. The consistency of these associations across diverse populations suggests that these metabolomic changes represent biological responses to dietary patterns regardless of population background. Upon integrating proteomic, metabolomic, and Mendelian randomization findings from the current stutdy, we identified several shared biological pathways through which diverse dietary patterns influence cardiometabolic health (**Figure 4**).

#### 1. The common biological pathway from healthy diets to a reduced risk of cardiometabolic disease, mediated through large HDL particles (LHDL) and apolipoprotein F (APOF) interactions

Greater adherence to healthy dietary patterns was linked to a reduced risk of cardiometabolic disease. This effect appears to be mediated by changes in large HDL particles and their associated APOF interactions. APOF, known as a lipid-transfer inhibitor protein, is predominantly carried on HDL and serves as a “metabolic switch” in HDL remodeling. By binding to HDL, APOF selectively inhibits CETP (cholesteryl ester transfer protein)-mediated transfer of cholesteryl esters from HDL to LDL, while favoring transfer to VLDL^33,34^. In our study, adherence to healthy dietary patterns consistently increased the concentration of LHDL, i.e., the HDL subclass most efficient at reverse cholesterol transport, which facilitates the delivery of APOF to sites of cholesterol efflux. At these sites, APOF’s inhibition of CETP fine-tunes HDL function, enhancing the removal of cholesterol from macrophages and reducing foam cell formation, key steps in preventing atherosclerosis and lowering cardiometabolic disease risk^33^.

#### 2. The common biological pathway from healthy diet to a reduced CMD risk, mediating through DNAJA2/Hsp40/EIF4G1-triglyceriderich lipoproteins interactions

DNAJA2 (DnaJ heat shock protein family (Hsp40) member A2) serves as a key co-chaperone for Hsp70 that guides nascent or stress-denatured proteins toward correct folding, prevents toxic aggregation, and preserves cellular proteostasis^35^. Beyond its classical role in protein quality control, Hsp40 co-chaperones also facilitate the early folding and lipidation of apolipoprotein B during VLDL assembly^36^, directly influencing the number and composition of triglyceride-rich- lipoproteins (TRL) in the bloodstream. These dysregulations contribute to endoplasmic reticulum (ER) stress, inflammation, thereby linking lipoprotein overproduction to broader metabolic dysfunction^37,38^. At the same time, ER stress activates selective translational programs through pathways such as eIF2α phosphorylation and ATF4 induction, where translation-initiation scaffolds such as EIF4G1 play a pivotal role^39^. Thus, the observed association of TRL with EIF4G1 may reflect an integrated chaperone-translation coupling mechanism; DNAJA2-mediated modulation of ApoB folding feeds back into EIF4G1 activity, reprogramming translation toward proteins required for ER homeostasis and lipid handling. Because dietary composition could influence VLDL-ApoB secretion and clearance, and thereby modulate postprandial triglyceride excursions, our findings suggest a pathway in which a healthy diet shapes lipoprotein metabolism and cellular stress responses through DNAJA2/Hsp40 engagement and EIF4G1-mediated translational reprogramming, ultimately impacting cardiometabolic risk.

#### 3. The biological pathway from healthy diet to a reduced CMD risk, mediating through CRHBP’s modulation of the HPA axis and consequent effects on triglyceride-rich lipoproteins

Corticotropin-releasing hormone binding protein (CRHBP) regulates hypothalamic-pituitary-adrenal (HPA)-axis activity by inactivating corticotropin-releasing hormone in the circulation, thereby limiting adrenocorticotropic hormone release and downstream cortisol production by the adrenal cortex^40^. Because cortisol drives hepatic triglyceride synthesis, enhances VLDL-ApoB100 assembly, and inhibits lipoprotein clearance, even modest alterations in CRHBP levels or binding affinity can shift VLDL particle number and triglyceride burden in the bloodstream^41^. Thus, CRHBP functions as an upstream gatekeeper of lipoprotein metabolism, and its dysregulation may promote dyslipidemia, insulin resistance, and broader cardiometabolic risk. Consistent with this mechanism, our study identifies a dietary influence, via overall diet quality (AHEI) and a plant-based pattern, on cardiometabolic health that is mediated through CRHBP’s modulation of the HPA axis and consequent effects on triglyceride-rich lipoproteins.

### Strengths and Limitations

Our study has several strengths. First, inclusion of four geographically and ethnically diverse cohorts enhances generalizability and enables identification of consistent biological signatures across populations. Second, integration of comprehensive multi-omics data provides a more complete picture of the biological pathways linking diet to cardiometabolic disease. Third, use of multiple dietary pattern scores captures distinct aspects of dietary quality and strengthens inferences about healthy dietary patterns more generally.

However, we acknowledge several limitations. First, the use of different dietary assessment methods across cohorts may introduce heterogeneity, though our standardization approaches mitigate this concern. Additionally, diets were only measured at baseline, and unmeasured diet changes over time could attenuate observed associations. Second, the cross-sectional nature of the omics measurements prevents establishing temporal relationships between dietary patterns, biomarker changes, and disease outcomes, required for mediation. Third, outcome definitions differed across cohorts (incident outcomes in UKB and NEO versus baseline phenotypes in GNHS and 10K), reducing direct comparability. For this reason, we emphasized consistency of effect direction rather than effect size when comparing prospective and cross-sectional results. Fourth, sample size limitations in the proteomics subsets resulted in wide confidence intervals for mediation estimates, indicating uncertainty in these results. Fifth, mediation estimates were sensitive to imprecision in direct effects with wide CIs that included the null, underscoring that these particular mediation results should be interpreted with caution. Sixth, ancestry differences between our cohorts and the largely European ancestry GWAS used to select genetic instruments mean that Mendelian randomization results are most directly applicable to European ancestry and may not generalize across ancestries. Also, a smaller number of instrumental variables available for proteins may account for the non-significant associations observed from proteins to metabolite levels.

## Conclusion

In this multi-cohort study integrating healthy dietary patterns, multi-omics profiles, and cardiometabolic outcomes across diverse populations, we demonstrate consistent associations between adherence to healthy dietary patterns and lower cardiometabolic disease risk. We identify replicable protein and metabolite signatures associated with dietary patterns and provide evidence that specific biomarkers, involved in lipid metabolism and immune function, mediate the association between diet and cardiometabolic disease. These findings enhance our understanding of the molecular mechanisms linking diet to cardiometabolic health across diverse populations and may inform the development of targeted dietary interventions and potential therapeutic strategies for the prevention of cardiometabolic disease.

## Methods

### Study design and cohorts

This analysis integrates data from four population-based cohort studies with diverse ethnic backgrounds and habitual diet: the NEO study^12^, UKB^13^, 10K study^14^, and GNHS^15^. All studies obtained ethical approval from their respective institutional ethics committees, and all participants provided written informed consent. This study adheres to the Strengthening the Reporting of Observational Studies in Epidemiology (STROBE) guidelines for reporting observational studies.

The NEO study is a population-based cohort study including 6,671 men and women aged between 45 and 65 years at baseline (2008-2012), with an oversampling of individuals with a BMI ≥27 kg/m². The UKB is a population-based prospective cohort of approximately 500,000 participants aged 40-69 years recruited across the UK between 2006 and 2010. The 10K study is a large-scale prospective longitudinal cohort established in Israel, including more than 10,000 participants aged 40-70 years at baseline. The GNHS is a longitudinal cohort study, which started in 2008, including approximately 4,048 healthy residents aged 40-80 years living in Guangzhou city (South China) for more than 5 years. Due to the short follow-up period in the 10K cohort (only 2 years) and the imprecise follow-up time in the GNHS cohort (only the year of diagnosis, without a specific date, was available), these two cohorts were analyzed cross-sectionally. In contrast, the NEO and UKB cohorts were analyzed prospectively. Detailed descriptions of each cohort are provided in the **Supplemental Methods**.

**Figure S1** presents a flow diagram of participant selection across the four cohorts. Because we used different analytical approaches (prospective or cross-sectional), exclusion criteria were adapted accordingly. In NEO and UKB cohorts, which were used for prospective analyses of disease incidence, we excluded participants if 1) they had type 2 diabetes or cardiovascular disease at baseline; 2) follow-up data were unavailable; 3) reported energy intake was implausible (<600 kcal/day or >5000 kcal/day); or 4) key variables were missing. After applying these exclusion criteria, the final study population included 4,838 participants from the NEO study (median follow-up 6.6 years, interquartile range [IQR] = 5.8-7.4), 54,024 participants from UKB (median follow-up 12.7 years, IQR = 12.4-13.1).

In contrast, we did not exclude participants with existing cardiometabolic diseases at baseline in the GNHS and 10K cohorts. Instead, prevalent cases of type 2 diabetes and dyslipidemia were identified based on clinical and biochemical criteria as described below. After applying the same additional exclusion criteria (implausible energy intake and missing variables), the final sample included 3,201 participants from GNHS and 9,616 from 10K.

### Dietary pattern scores

Dietary intake was assessed using food frequency questionnaires (FFQ) in NEO and GNHS, 14-day dietary logging in 10K, and Oxford WebQ (online 24-hour dietary questionnaire) in UKB^16^. Previous studies showed the comparable performance of Oxford WebQ with traditional interviewer-based 24-hour recall^17^, and FFQ with 24-hour recalls and multiple-day dietary records^18^. For each cohort, five dietary pattern scores were calculated: AMED^6^, hPDI^7,8^, DII^9^, EDIH^10^, and AHEI^11^. To ensure comparability of dietary pattern scores across cohorts, we followed the original literature’s recommendation for standardized food group categorizations and scoring algorithms. Detailed descriptions of the components and scoring method are presented in **Supplemental Methods** and **Tables S13-S17**.

Additionally, we used energy-adjusted values and cohort-specific distributions for all dietary scores to minimize systematic differences due to dietary assessment instruments. Energy-adjusted dietary pattern scores were calculated using the sex-specific residual method, regressing dietary pattern scores on total energy intake (kcal/day). Winsorization was applied based on the 0.5th and 99.5th percentiles to minimize the influence of extreme values. Scores were standardized based on SD within each cohort. For consistent interpretation, the EDIH and DII scores were reversed so that higher values indicated healthier diets across all scores. **Figure S3** shows the relationship between energy-adjusted dietary pattern scores in each cohort.

### Ascertainment of outcomes

The primary outcome was the incidence of cardiometabolic diseases, defined as the occurrence of either cardiovascular disease or type 2 diabetes. Follow-up time was calculated from baseline to the first occurrence of either disease diagnosis, death, loss to follow-up, or the end of follow-up, whichever came first. Secondary outcomes included the incidence of each separate disease.

In the NEO study, cases were identified using electronic health records maintained by general practitioners (GPs)^19^. Type 2 diabetes diagnoses were based on the International Classification of Primary Care (ICPC) codes T90 (diabetes mellitus) or T90.02 (diabetes mellitus type 2) in combination with prescriptions for medications under the Anatomical Therapeutic Chemical (ATC) code A10. New diagnoses of cardiovascular disease were determined by identifying cases of myocardial infarction (ICPC: K75, K76.02), transient ischemic attack (K89), and cerebrovascular accident (K90).

In UKB, cases were identified using primary care data, hospital inpatient records, death register entries, and self-reported medical conditions. Type 2 diabetes was based on ICD-10 code E11 (non-insulin-dependent diabetes mellitus), while cardiovascular disease was identified using ICD-10 codes I21-I23 (myocardial infarction), I63 (ischemic stroke), and I64 (stroke not specified as hemorrhage or infarction). For both NEO and UKB, follow-up time was defined as described above, with the end of data collection on December 31, 2018, for NEO and October 31, 2022, for UKB.

In GNHS and 10K, we used prevalent cases at baseline. In GNHS, type 2 diabetes was defined as fasting glucose ≥ 7.0 mmol/L (126 mg/dL), or HbA1c ≥ 6.5% (48 mmol/mol), or with diabetes medication use. In 10K, type 2 diabetes was identified based on continuous glucose monitoring (CGM) with glucose monitoring index ≥6.5%, or mean glucose >154 mg/dL, or >20% time above 180 mg/dL, or self-reported use of medications for type 2 diabetes. Dyslipidemia was assessed in both cohorts through total cholesterol ≥6.2 mmol/L, triglycerides ≥2.3 mmol/L, LDL cholesterol (LDL-C) ≥4.1 mmol/L, HDL cholesterol (HDL-C) <1.0 mmol/L, or lipid-lowering medications.

Given these methodological differences across cohorts, we primarily focused on the consistency of effect directions rather than the magnitude of associations when comparing results between prospective analyses (NEO, UKB) and cross-sectional analyses (GNHS, 10K).

### Proteomic and metabolomic profiling

Only UKB, NEO, and GNHS have data on proteomics. Among the three cohorts, 3,197 unique proteins were measured (UKB: 2,923, NEO: 363, GNHS: 328) in total. There was some overlap between the proteins measured in each cohort, with 3 proteins measured in all three cohorts, 358 in both NEO and UKB, and 53 in both UKB and GNHS. In UKB, plasma proteins were quantified using the Olink Explore 3072 platform^20^. In NEO, proteins in plasma were quantified using the Olink Explore 384 Inflammation panel^21^, according to the standard Olink protocol. In GNHS, baseline serum proteins were characterized using mass spectrometry (MS)-based proteomics, as described previously^22^. Standard quality control procedures were followed according to each platform’s specifications. In GNHS, missing protein levels were imputed using half of the minimum value for each protein, and batch effects were corrected using the R package ‘proBatch’.

Across all four cohorts, metabolomic measurements included 326 unique metabolites (UKB: 168, NEO: 148, GNHS: 164, 10K: 165), with 12 metabolites measured in all four cohorts and 127 measured in UKB, NEO, and 10K. UKB, NEO, and 10K measured metabolite profiles using the Nightingale Health ^1^H nuclear magnetic resonance platform^23^. In GNHS, targeted metabolomics profiling was performed using the Q300 Metabolite Assay Kit with serum samples processed via UPLC-MS/MS^24^. In all cohorts, participants with >70% missing metabolite measurements were excluded, and metabolites with >40% missing values were removed. The remaining missing metabolite levels were imputed with half the minimum observed value for each metabolite. Rank-based inverse normalization was applied to transform the distribution of each metabolite. Detailed descriptions of omics data acquisition and quality control are provided in the **Supplemental Methods**.

### Statistical analysis

Baseline characteristics of study populations were summarized as means with SD for continuous variables or numbers with percentages (%) for categorical variables.

#### Associations between dietary patterns and clinical outcomes

We examined associations between dietary patterns and cardiometabolic outcomes using different regression approaches based on cohort follow-up data availability. For the prospective analyses in UKB and NEO, we applied Cox proportional hazard regression models to assess associations between individual dietary patterns and incident cardiometabolic disease, type 2 diabetes, and cardiovascular disease. For GNHS and 10K with limitations in their follow-up data, we used logistic regression to evaluate associations between dietary patterns and baseline cardiometabolic phenotypes (i.e., prevalent cases).

Models were adjusted for age, sex, smoking status, physical activity, vitamin use, hormone medication use, socioeconomic status reflected by either education levels (NEO, 10K, and GNHS) or Townsend deprivation index (UKB), and sleep duration. We used cohort-specific definitions for some of the covariates, as detailed in the **Supplemental Methods**. Alcohol consumption was additionally adjusted for in the association analyses when hPDI was used as the exposure. Dietary pattern scores were analyzed as continuous variables (per SD). As the sensitivity analyses, models were adjusted for BMI as a continuous variable, given its potential role as a mediator rather than a confounder. Additional adjustments were made for family history of type 2 diabetes or cardiovascular disease and nonsteroidal anti-inflammatory drugs (NSAID) usage in UKB, NEO, and 10K, but not in GNHS due to the lack of data.

#### Mediation analyses using multi-omics data

We assessed associations between dietary patterns and omics biomarkers using linear regression models with standardized variables, expressing results as SD difference in metabolite/protein levels per SD in a dietary pattern score. For associations between omics biomarkers and disease outcomes, we employed Cox proportional hazard regression models in UKB and NEO, and logistic regression models in GNHS and 10K, mirroring our approach for dietary pattern analyses. All analyses were adjusted for the confounders mentioned above. The Benjamini-Hochberg procedure was used to correct for multiple testing in UKB (q < 0.05), while nominal p-values (p < 0.05) were used for replication in the other cohorts.

For mediation analysis, we first addressed the high dimensionality and correlation structure of metabolites using the identification of principal variables (iPVs) approach^25^. This method identifies representative metabolites from clusters of highly correlated features using a multi-step process: (1) generating a correlation matrix using Spearman’s rank correlation; (2) converting correlations to distances; (3) performing hierarchical clustering with complete linkage; (4) cutting the dendrogram at height 0.5; and (5) selecting one representative metabolite from each cluster using principal variable analysis. This approach reduces dimensionality while retaining biologically meaningful signals and minimizing multicollinearity in mediation analyses.

Using the identified principal markers, we performed mediation analyses. For each dietary pattern-disease combination, we estimated: (1) the total effect (dietary pattern on disease without mediators); (2) the direct effect (dietary pattern on disease adjusting for mediators); and (3) the indirect effect (which measures how much of the dietary pattern’s effect occurs through changes in the mediator). The mediated proportion was calculated as the ratio of indirect effect to total effect. Bootstrapping with 50 resamples was used to derive 95% confidence intervals for all mediation parameters.

To meet the assumptions of mediation analysis, the exposures (i.e., dietary patterns) must be associated with both the outcomes (i.e., cardiometabolic disease incidence) and the mediators (i.e., omic markers). However, in the NEO and GNHS, some of these required associations were not observed, which may in part reflect limited sample sizes. As a result, the mediation analysis could not reliably estimate the proportion mediated in these cohorts. Therefore, we chose to report mediation analysis results only from the UK Biobank, although part of the associations (e.g., exposure-mediator or exposure-outcome) were also replicated in either NEO or GNHS.

#### Mendelian randomization analysis

To more comprehensively characterize the underlying biological pathways, bidirectional Mendelian randomization analyses were conducted to explore potential causal associations between circulating metabolite and protein levels by integrating multiple omics layers. Analyses were performed exclusively on pairs where both the metabolite and protein were associated with the same dietary pattern and clinical outcome. Genome-wide significant genetic variants for each metabolite and protein were selected from large-scale GWAS performed in the European population^26,27^.

The primary Mendelian randomization estimate was generated via the inverse variance weighted (IVW) method, with sensitivity analyses using MR Egger regression and the Weighted Median estimator. We addressed the three core instrumental variable assumptions. First, for relevance, we selected genetic variants strongly associated with each exposure (p < 5×10⁻⁸) and calculated F-statistics to ensure instrument strength (all F>10). Second, for independence, we applied MR-Egger regression to test for directional pleiotropy. Third, for the exclusion restriction, we employed multiple sensitivity analyses (weighted median and MR-Egger) to detect potential violations. Heterogeneity among instruments was assessed using Cochran’s Q statistic. A Bonferroni correction was applied to account for multiple testing (significance threshold: 0.05/59).

### Software and data availability

All statistical analyses were performed using R version 4.1.0. iPVs were implemented using iPVS package^25^. Mediation analyses were implemented using CMAverse package^28^. MR analyses were implemented using the TwoSampleMR package^29,30^. Study protocols, analytic code, and summary statistics will be made available upon reasonable request to the corresponding author. Original data are subject to access restrictions according to each cohort’s data sharing policies.

## Data Availability

Original data are subject to access restrictions according to each cohort's data sharing policies.

## Acknowledgment

We thank all participants of the NEO study and all participating general practitioners for inviting eligible participants. We also express our gratitude to Pat van Beelen and all research nurses for the data collection, Petra Noordijk and her team for the laboratory management, and Ingeborg de Jonge for data management. We thank Shirley Bao for her work on the NEO Olink data during her internship. The authors wish to acknowledge all participants of the 10K and all colleagues who contributed to this work. We thank all participants of the Guangzhou Nutrition and Health Study for their invaluable contribution, as well as all team members involved in the cohort study. This research has been conducted using the UK Biobank Resource under Application Number 242500, and we thank the UK Biobank participants and coordinators for providing access to these data.

## Funding

The NEO study is supported by the participating departments, the Division and the Board of Directors of the Leiden University Medical Centre, and by the Leiden University, Research Profile Area ‘Vascular and Regenerative Medicine’. R.L-G is supported by JPI HDHL NUTRIMMUNE DIYUFOOD project. K.D is supported by China Scholarship Council (No. 202206210140). All the funders didn’t participant in the process of the whole study.

## Conflict of Interest

All authors state that they have no conflict of interest.

## Author contribution

J.H, K.D, R.L, and A.v.H.V contributed to the conceptualization and study design of the current study. J.H, K.D, and Z.H performed statistical analyses. J.H drafted the manuscript. R.d.M and F.R.R conducted the study design and data collection of the NEO study. N.G, and E.S conducted the study design and data collection of the 10K study. Z.Z, J.Z, and Y.C conducted the study design and data collection of the GNHS study. All authors reviewed the manuscript and approved the final version of the manuscript.

## Notes

### Competing Interest Statement

The authors have declared no competing interest.

### Author Declarations

All studies obtained ethical approval from their respective institutional ethics committees, and all participants provided written informed consent. All the four datasets that we used in the manuscript are de-identified.

