## Supplemental Material for "Multi-Omics characterization of biological pathways linking healthy dietary patterns to cardiometabolic disease risk across diverse populations"

Ruifang Li-Gao

Department of Clinical Epidemiology, Leiden University Medical Center, Leiden, the Netherlands.

Albinusdreef 2, 2333 ZA Leiden, the Netherlands

### Supplemental Methods

#### Description of study cohorts

##### Netherlands Epidemiology of Obesity study (NEO)

The NEO study is a population-based cohort study including 6671 men and women aged between 45 and 65 years at baseline (2008-2012), with an oversampling of individuals with a self-reported body mass index (BMI) of 27 kg/m^2^ or higher. The study was approved by the Medical Ethical Committee of the Leiden University Medical Center (LUMC), Leiden, the Netherlands. All participants gave their written informed consent. The study design has been described in detail previously (1). Briefly, people with a BMI of 27 kg/m2 or higher living in the greater area of Leiden in the west of the Netherlands were invited to participate in the NEO study. Additionally, people living in the neighboring municipality Leiderdorp were invited regardless of BMI, representing a reference distribution of BMI similar to the general Dutch population. Participants were invited to a baseline visit at NEO study centre of the LUMC after an overnight fast. Prior to this study visit, participants collected their urine over 24 h and completed a general questionnaire at home to report demographic, lifestyle and clinical information, additionally they completed questionnaires on diet and physical activity. The participants were asked to bring all medication they were using to the study visit. At the baseline visit several measurements were performed including anthropometry and blood pressure, and blood samples were drawn. Participants were followed by the electronic patient records of general practitioners for clinical endpoints including type 2 diabetes, cardiovascular disease, and mortality. Habitual dietary intake of all participants was estimated through use of a self-administered, semiquantitative 125-item food frequency questionnaire (FFQ) (2-4). In this FFQ, participants were asked about the frequency of intake of foods during the past month (times per day, times per week, times per month, or never). In addition, the serving size for each food was estimated. Dietary intake of nutrients and total energy was estimated using the Dutch Food Composition Table (NEVO-2011).

### 10K

The 10K study, a large-scale prospective longitudinal cohort that was established in Israel (5). The 10K study aims to identify novel molecular markers with a diagnostic, prognostic and therapeutic value. The study is expected to include 10,000 participants aged between 40 and 70 years from the general population of Israel, and by the end of 2021, the baseline inclusion was achieved. Information collected at baseline includes medical history, lifestyle and nutritional habits, vital signs, anthropometrics, blood tests results, electrocardiography (ECG), ankle-brachial pressure index (ABI), liver ultrasound (US) for liver diseases and dual-energy X-ray absorptiometry (DXA) tests for bone mineral density as well as total body composition. In addition to extensive omics profiles on genome, transcriptome, proteome, gut and oral microbiome, metabolome and immune system profiling, continuous measurements are also available, including glucose levels measured using a continuous glucose monitoring (CGM) device for two weeks coupled with self-detailed logging of daily activities (such as food intake, sleep times and physical activity) and sleep monitoring by a home sleep apnea test (HSAT) device for 3 nights. The study plans to have follow-up visits every year for a total of 25 years. The linkage to national disease registries is also ongoing.

##### Guangzhou Nutrition and Health study (GNHS)

Guangzhou Nutrition and Health Study (GNHS) is a robust longitudinal cohort study that began in 2008 and conducts follow-up assessments approximately every 3 years (6). About 4048 apparently healthy residents, living in Guangzhou city (South China) for more than 5 years, aged 40-80 years, were recruited between 2008 and 2013. At each visit, detailed sociodemographic data, health-related lifestyle factors, and medical history information were obtained by a structured questionnaire administered in face-to-face interviews. A 19-item physical activity questionnaire was used to assess daily physical activity and the metabolic equivalent intensity (7). The participants were asked to recall their usual dietary consumption using a 79-item food frequency questionnaire (FFQ) (8). The dietary daily intakes of total energy and nutrients were calculated according to the Chinese Food Composition Table, 2002 (9). Overnight fasting blood samples were collected, separated, and stored at -80°C till tests.

##### UK Biobank (UKB)

The UK Biobank is a population-based prospective cohort of approximately 500,000 participants aged 40-69 years recruited across the UK between 2006 and 2010. Participants underwent comprehensive baseline assessments, including biological measurements, lifestyle indicators, and biomarkers from blood and urine samples. Follow-up was conducted through linkage to health and medical records (10).

#### Dietary assessment

##### Alternate Mediterranean Diet (AMED)

The AMED score comprised nine components: fruits, vegetables (excluding potatoes), whole grains, nuts, legumes, fish, red and processed meat, the ratio of monounsaturated fat to saturated fat, and alcohol (11). For most components, participants with intakes above the population median received 1 point; those below received 0. For red and processed meat, the scoring was reversed. Alcohol intake scoring was based on sex-specific median values, except in GNHS where alcohol intake ≤15g received 1 point based on Chinese Dietary Guidelines (12). The detailed food components used are shown in **Table S1.**

##### Healthful Plant-Based Diet Index (hPDI)

The hPDI categorized 18 food groups into three types: healthy (e.g., whole grains, fruits, vegetables, nuts, legumes, vegetable oils, and tea/coffee), less healthy (e.g., fruit juice, refined grains, potatoes, sugar-sweetened beverages, and sweets/desserts), and animal-based (e.g., animal fat, dairy, eggs, fish/seafood, meat, miscellaneous animal-based foods) (13, 14) . Due to data limitations in the GNHS, animal fat and miscellaneous animal-based foods were excluded. Quintiles of food group intake (g/day) were used to assign points: 5 points for the highest quintile of healthy food intake and the lowest quintile of less healthy or animal-based food intake, with 1 point assigned otherwise. The detailed food components used are shown in **Table S2.**

##### Dietary Inflammation Index (DII)

Shivappa et al. derived food-specific inflammatory effect scores (DII) based on 1,934 studies that examined dietary effects on inflammatory biomarkers, including interleukin‐1β, interleukin‐4, interleukin‐10, tumor necrosis factor-alpha, interleukin‐6, and C-reactive protein (15). The DII quantifies dietary inflammatory potential using various food categories. **Table S3** shows the list of food categories used in each cohort. For dietary intake in each cohort under study, the representative world databases that provides a mean and standard deviation for consumption of each food component in the global population. Food intake of individual was expressed as a relative to the “standard global mean” as a Z-score: (diet intake – standard global mean)/global SD. To minimize the effect of right-skewing, this value is converted to a percentile score. To further achieve a symmetrical distribution with values centered on 0 (null) and bounded between −1 (maximally anti- inflammatory) and +1 (maximally pro-inflammatory), each percentile score is doubled and then ‘1’ is subtracted. The centered percentile value for each dietary component is multiplied by its respective ‘overall dietary component-specific inflammatory effect score’ to obtain the ‘dietary component specific DII score’. Finally, all the dietary component-specific inflammatory effect scores are summed up to indicate overall DII score for an individual.

##### Empirical Dietary Index for Hyperinsulinemia (EDIH)

The EDIH was based on 18 food groups most predictive of hyperinsulinemia: red meats, low-energy beverages, cream soup, processed meats, margarine, poultry, butter, potato fries, fish, high-energy beverages, tomato, low-fat dairy, eggs, wine, coffee, fruit, high-fat dairy, vegetables (16). Due to data limitations in the NEO study, low-energy beverages, cream soup, poultry, and tomatoes were excluded. Due to data limitations in the GNHS, low energy beverages, cream soup, margarine, butter, and french fries were excluded. In 10K, due to the lack of information, margarine was excluded when calculated EDIH. Food groups were analyzed as grams per day. The EDIH score was calculated as the weighted sum of food-specific intake scores. **Table S4** shows the detailed weights for each food component.

##### Alternate Healthy Eating Index (AHEI)

The AHEI was calculated based on the intake of 11 food and nutrient groups: vegetables, fruits, whole grains, sugar-sweetened beverages and fruit juice, nuts and legumes, red and processed meat, alcohol, sodium, trans fats, long-chain fatty acids, and polyunsaturated fatty acids (17). Scores ranged from 0–10 points per group, proportional to daily intake. Sex-specific criteria were applied to score alcohol intake. Sodium was excluded due to the data being unavailable in the NEO study. Detailed food components used for calculation are shown in **Table S5.**

#### Metabolomics data

##### Netherlands Epidemiology of Obesity study (NEO)

NEO participants had plasma-derived metabolite profiles measured using the [Nightingale](https://www.sciencedirect.com/topics/biochemistry-genetics-and-molecular-biology/luscinia) Health (Helsinki, Finland) ^1^H nuclear magnetic resonance (NMR) platform (18). For each participant, both their fasting and 150-min postprandial samples were assayed. We note here that metabolites are commonly defined as biological molecules of <1.5 kDa in size and that many of the molecules (lipids) assayed in this study are larger than this threshold. For simplicity, we will refer to them all as metabolites. At the time of sampling, this [metabolomics](https://www.sciencedirect.com/topics/agricultural-and-biological-sciences/metabolomics) platform provided 229 measurements for 149 metabolites, including 80 derived ratio measurements from 14 substance classes: [amino acids](https://www.sciencedirect.com/topics/agricultural-and-biological-sciences/amino-acid) (*n* = 8), [apolipoproteins](https://www.sciencedirect.com/topics/biochemistry-genetics-and-molecular-biology/apolipoprotein) (*n* = 3), cholesterol (*n* = 9), fatty acids (*n* = 11), fatty acids ratios (*n* = 8), fluid balance (*n* = 2), [glycerides](https://www.sciencedirect.com/topics/pharmacology-toxicology-and-pharmaceutical-science/acylglycerol) and [phospholipids](https://www.sciencedirect.com/topics/pharmacology-toxicology-and-pharmaceutical-science/phospholipid) (*n* = 9), glycerides and phospholipid ratios (*n* = 2), glycolysis-related metabolites (*n* = 3), inflammation (*n* = 1), [ketone bodies](https://www.sciencedirect.com/topics/pharmacology-toxicology-and-pharmaceutical-science/ketone-bodies) (*n* = 2), [lipoprotein](https://www.sciencedirect.com/topics/pharmacology-toxicology-and-pharmaceutical-science/lipoprotein) particle size (*n* = 3), [lipoprotein](https://www.sciencedirect.com/topics/biochemistry-genetics-and-molecular-biology/lipoprotein) subclasses (*n* = 98), and [lipoprotein](https://www.sciencedirect.com/topics/agricultural-and-biological-sciences/lipoprotein) subclass ratios (*n* = 70). For the current study, we excluded ratio measurement.

### 10K

The metabolomics data in 10K were generated from Nightingale Health's metabolic profiling of blood serum samples with an extensive array of metabolites, encapsulating lipids, fatty acids, amino acids, and various low-molecular-weight metabolites. Utilizing NMR technology, the dataset unveils a spectrum of 250 biomarkers, 39 of which are clinically validated, offering a profound insight into the metabolic mechanisms underlying health and disease. Specially, after collecting blood samples from participants during clinic visits, the blood samples are immediately placed in tubes and processed to separate the serum. These serum samples are then stored in plates and kept in the freezer to maintain their integrity until shipment. Upon arrival at Nightingale Health's laboratories, the serum samples are once again frozen. When ready for analysis, the samples are thawed and undergo a series of preparatory steps based on the standard instructions.

##### Guangzhou Nutrition and Health study (GNHS)

Targeted metabolomics profiling for baseline serum samples was performed using the Q300 Metabolite Assay Kit (Human Metabolomics Institute, Inc., Shenzhen, Guangdong, China), adapted from published methods (19). Previously published literature provides the details (20). Briefly, serum extracts (25 μL) were processed in 96-well plates using a Biomek 4000 workstation (Beckman Coulter, Brea, CA, USA) and mixed with internal standards. Following centrifugation (Allegra X-15R, Beckman Coulter, Indianapolis, IN, USA), the supernatants underwent derivatization at 30°C for 60 minutes. Additional internal standards were added after methanol dilution, and serial dilutions of stock standards were prepared. Metabolites in serum samples were quantified using UPLC-MS/MS (ACQUITY UPLC-Xevo TQ-S, Waters, Milford, MA, USA), operated with MassLynx 4.1 software. Chromatographic separation was performed using an ACQUITY BEH C18 column (1.7 μm, 100 mm × 2.1 mm, Waters, Milford, MA, USA). Peak integration, calibration, and quantification were conducted using TMBQ 177 (v1.0, Human Metabolomics Institute, Shenzhen, China). Standard curves were validated for each sample batch to ensure quantification accuracy, with mixed standards and biological samples included in every batch to monitor detection quality. Ultimately, 196 serum metabolites were accurately quantified, with fewer than 40% missing values. Metabolites with a coefficient of variation (CV) greater than 25% based on quality control (QC) samples were excluded, leaving 165 metabolites for further analysis, with the CV values of QC samples ranging from 2.5% to 23.7%.

##### UK Biobank (UKB)

Plasma metabolites profiling in UKB was conducted by Nightingale Health using a 500 MHz NMR spectroscopy platform on EDTA plasma samples (21). Blood samples, collected at 22 assessment centers, were processed into 96‑well plates and shipped frozen to Finland, where they were thawed, centrifuged, and prepared for analysis. Two spectra were recorded for each sample: a pre-saturated proton spectrum capturing signals from proteins and lipoprotein lipids, and a T₂-relaxation-filtered spectrum enhancing the detection of low-molecular-weight metabolites. This approach quantified 249 metabolic measures including 168 absolute concentrations and 81 ratios with robust quality control procedures in place.

#### Proteomics data

##### Netherlands Epidemiology of Obesity study (NEO)

Proteins in the collected plasma at baseline from a random subset of NEO participants (n = 1545) were quantified using the Olink Explore 384 Inflammation panel using proximity extension assay. Three proteins (i.e., BCL2L11, BID, and MGLL) did not meet the manufacturer’s batch release quality control criteria and therefore are not included in the analyses. As a result, 365 proteins were measured in the NEO study.

##### Guangzhou Nutrition and Health study (GNHS)

Baseline serum proteins were characterized and quantified using MS-based proteomics (22, 23). Peptide samples were derived from serum and analyzed through an Eksigent NanoLC 400 System coupled with a TripleTOF 5600 system (SCIEX) for SWATH-MS. Each sample underwent two to three runs employing a 20-minute DIA-MS method, as previously detailed (24). The resulting MS files were processed using DIA-NN software (v1.8) and matched to a spectral library containing 5,102 peptides and 819 unique proteins from the Swiss-Prot *Homo sapiens* database (23, 25). After data cleaning and filtering for inclusion, we obtained a proteomic matrix comprising 413 proteins. A detailed description of the data cleaning steps has been published previously (23). The dataset exhibited strong consistency and reproducibility, evidenced by median Pearson correlation coefficients of ≥0.93 across pooled serum samples, biological replicates, and technical replicates (23). We excluded proteins with more than 40% missing values, resulting in a final dataset of 328 proteins. Missing protein levels were imputed using half of the minimum value for each protein. Batch effects were corrected using the R package 'proBatch'. Prior to batch effect correction, proteomic data underwent log transformation and median normalization.

##### UK Biobank (UKB)

Plasma samples were initially collected from UK Biobank participants and subsequently stored at ultra-low temperatures (−80 °C and in liquid nitrogen) to maintain protein integrity. For proteomic analysis, blood samples from a randomly selected subset of participants (n = 54,967) were shipped to a specialized analysis facility in Sweden. Proteins in the collected plasma were quantified using the Olink Explore 3072 platform in March 2023. This platform measured 2,923 analyte targets, which correspond to 2,941 unique proteins, spanning the panels designated as Cardiometabolic I and II, Inflammation I and II, Neurology I and II, and Oncology I and II. A comprehensive description of participant selection and sample handling protocols is available in the previous literature (26).

#### Covariates data

##### Netherlands Epidemiology of Obesity study (NEO)

In the NEO study, participants reported their age, sex, smoking status, and family and personal medical history in the baseline questionnaire. Smoking status was categorized as current, former, or never smoker. Participants also reported the frequency and duration of their physical activity, which included walking, cycling, gardening, home repair, and sports, during leisure time over the past four weeks, using the Short Questionnaire to Assess Health-enhancing Activity (SQUASH), expressed in MET-hours per week. Vitamin intake was reported as either supplements or medications. In women, we classified the use of contraceptives and hormone replacement therapy as either current or past/never users. Habitual dietary intake was assessed using a self-administered, semiquantitative 125-item food frequency questionnaire. Nutrient and total energy intake were estimated using the Dutch Food Composition Table (NEVO-2011). Educational attainment was reported in ten categories based on the Dutch education system and classified into high (including higher vocational school, university, and postgraduate education) versus low education (reference). Sleep duration was measured with the Pittsburgh Sleep Quality Index (PSQI) questionnaire.

### 10K

In the 10K cohort, a subset of participants filled in the questionnaire at baseline visit. Age, sex, height and weight were recorded. BMI was calculated as body weight (kg) divided by square of height. The questionnaire followed the structure and content of the sociodemographic, lifestyle and environment and psychosocial factors questionnaires from the UK Biobank (27). The questionnaires included multiple aspects, such as smoking status (categorized as current, former and never smoker), physical activity (expressed in MET-hours/week, including walking, moderate and vigorous activity), education status (high: professional-high-school, practical engineer, electrician, Bachelor, Master and Doctorate and low: less than 12 years of schooling), sleep duration (hours/day), family history of type 2 diabetes and cardiovascular disease (yes/no). Alcohol intake (grams/day) was derived from the real-time self-logging diet data. Previous medication use was based on self-reports relating to if the medication was taken between the study enrollments. Self-reported medications are coded to Anatomical Therapeutic Chemical (ATC) codes from the WHO collaborating center for Drug Statistics Methodology. In details, 1) vitamin usage (yes/no) was decided based on ATC codes starting with A11*; 2) hormone usage (yes/no) was based on ATC codes starting with G03*, H01*, H02*, H03*, H04* and H05*; 3) Aspirin and other non-steroidal anti-inflammatory (NSAID) drug usage (yes/no) was based on ATC codes starting with B01*, M01* and N02*.

##### Guangzhou Nutrition and Health study (GNHS)

In the GNHS, weight (kg) was measured with a digital scale while participants wore light clothing and no shoes, and height (m) was measured using a stadiometer. Body mass index (BMI) was calculated as weight divided by height squared (kg/m²). Sociodemographic and health-related lifestyle data were collected via a structured questionnaire administered through face-to-face interviews at baseline. Smoking habit was reported as yes (if more than 5 packs were smoked in lifetime) vs. no (if 5 packs or less were smoked). Drinking status was reported as yes (if there was regular drinking, defined as at least once a week for 6 consecutive months, in the past year) vs. no (if there was no regular drinking or only occasional drinking). Vitamin use was considered "yes" if the individual had taken vitamins more than 30 times in the past year; otherwise, it was "no". The use of hormone medication (yes vs. no), sleep duration (in hours and minutes, converted to hours), and educational level (middle school or lower, high school or vocational college, or university or above) were all self-reported. Participants’ physical activity was assessed using a 19-item physical activity questionnaire, which included exercise, leisure activities, housework, occupational tasks, and other daily activities, along with their metabolic equivalent (MET) hours per day (7). Participants’ usual dietary intake was recalled using a 79-item food frequency questionnaire (FFQ), and daily total energy intake was calculated according to the 2002 Chinese Food Composition Table, as previously described (8, 9).

##### UK Biobank (UKB)

In the UKB, participants self-reported their age, sex, ethnicity, smoking status, sleep duration, and physical activity. Smoking status was categorized as current, former, or never smoker. Physical activity was assessed using a short form of the International Physical Activity Questionnaire and expressed in MET-hours per week for all activities, including walking, moderate, and vigorous activity. Vitamin and mineral supplement use was based on whether participants reported consuming any of 21 individual supplements the day before. Although this information was also collected at baseline, response rates were low at that time, and none of the participants in our study population provided data at baseline. Hormone medication use was assessed through the question: “Do you regularly take any of the following medications?”. Dietary intake was evaluated using the Oxford WebQ, a web-based 24-hour recall questionnaire, and total energy intake was calculated accordingly (Perez-Cornago et al., Eur J Nutr, 2021). Socioeconomic status was estimated using the Townsend deprivation index, which was assigned to participants based on the output area corresponding to their postcode. Sleep duration was assessed by asking, "About how many hours sleep do you get in every 24 hours? (Please include naps)."

### Supplemental Figure

#### Supplemental Figure 1. Flowchart of Inclusion and Exclusion Criteria for Each Individual Study

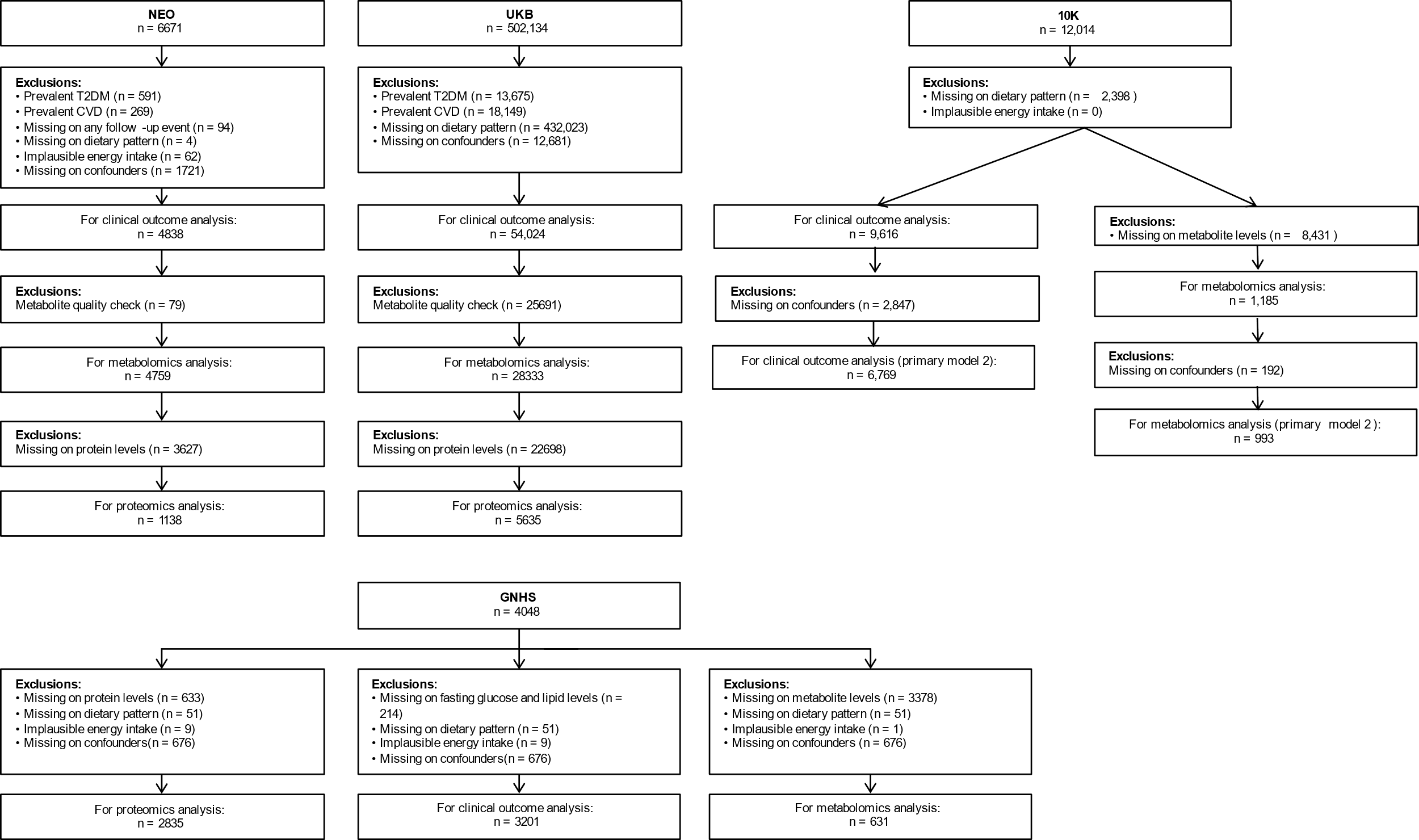

Selection process for each cohort showing exclusions for prevalent disease, missing data, and implausible energy intake (<600 or >5,000 kcal/day).

#### Supplemental Figure 2. Diet-related metabolites associated with cardiometabolic disease outcomes

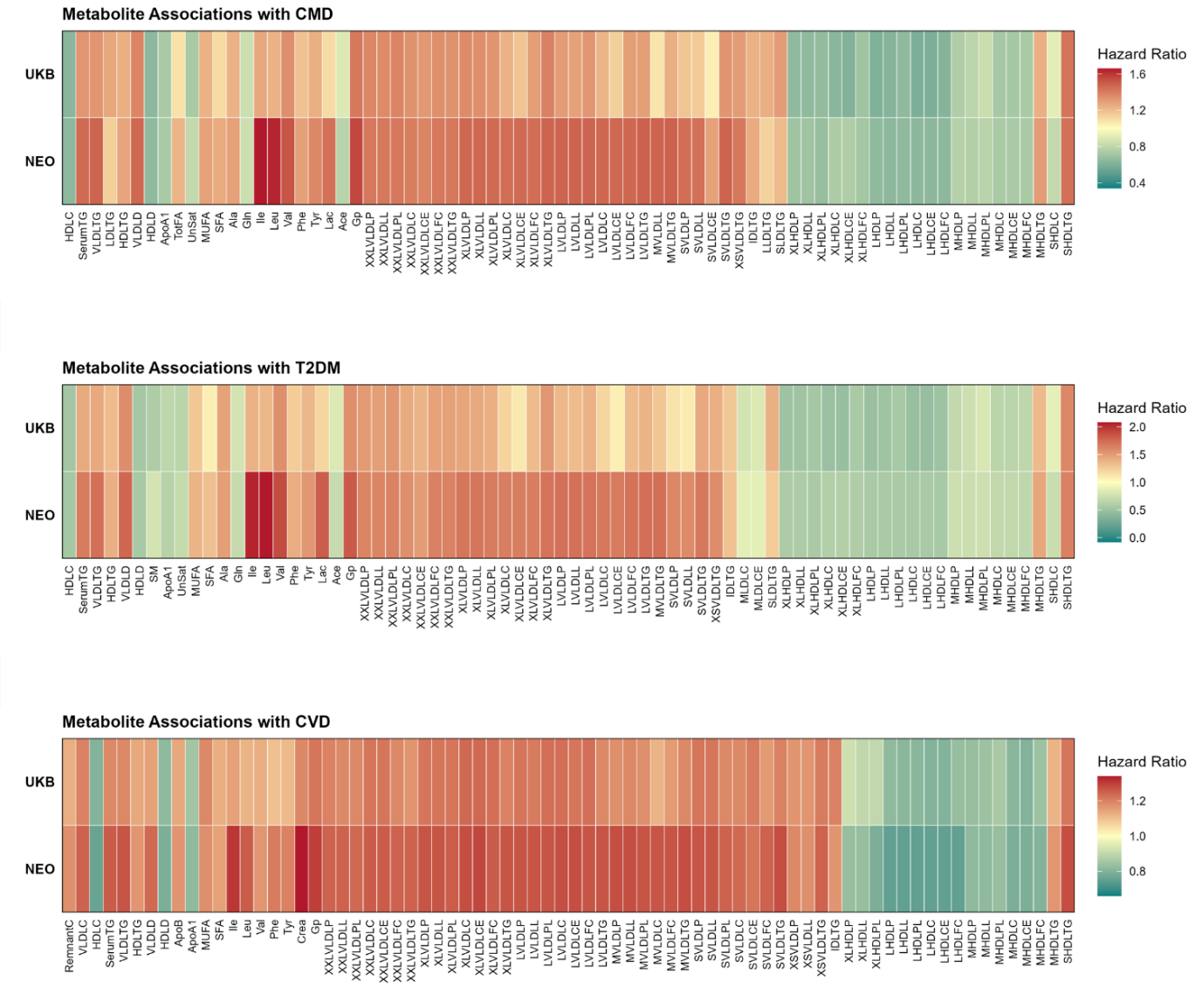

Heatmap showing associations between dietary pattern-related metabolites and cardiometabolic outcomes. Colors represent hazard ratios with blue indicating protective associations and red indicating harmful associations. Only metabolites significantly associated with both dietary patterns and outcomes are shown.

#### Supplemental Figure 3. Correlation plots between dietary patterns in each cohort

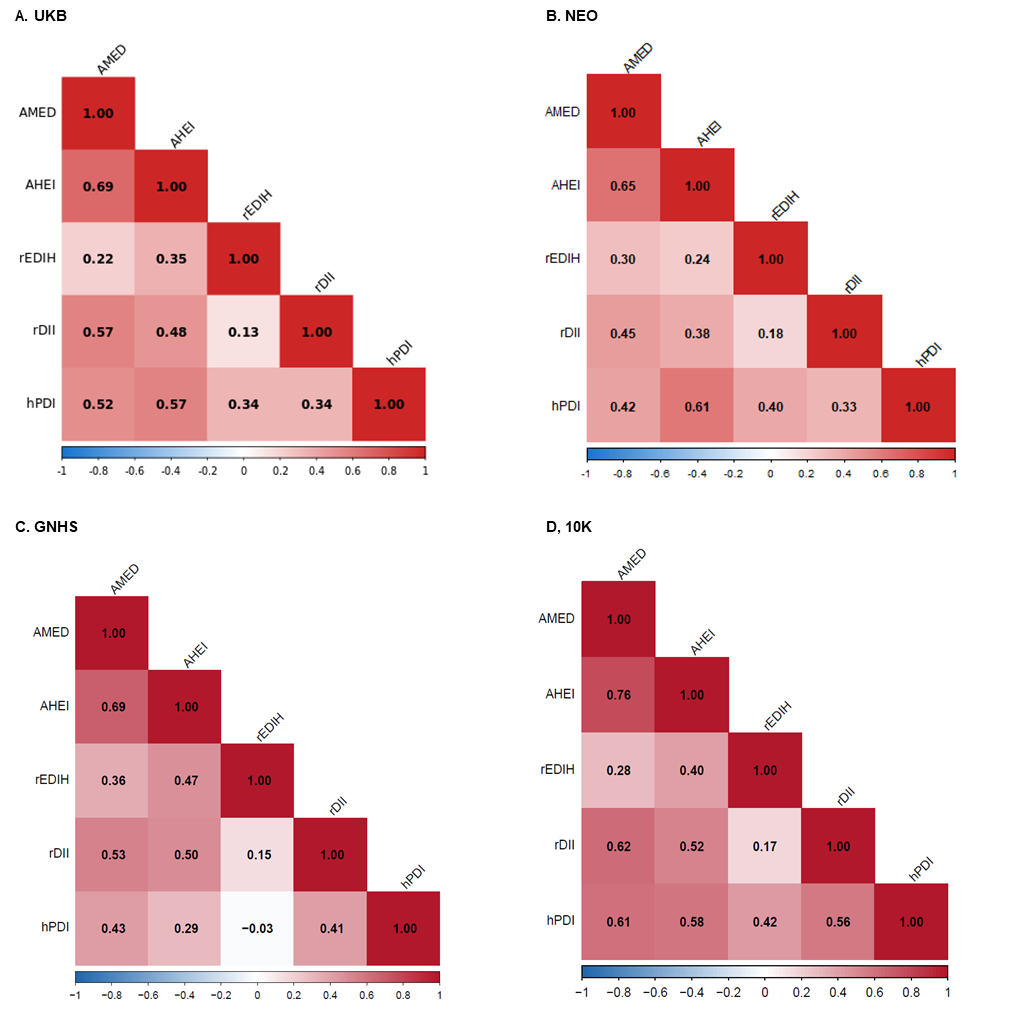

Spearman correlation coefficients between the five dietary pattern scores in each cohort.

### Supplemental Table

The tables below are provided in a separate file.

#### Table S1. Associations between dietary pattern scores and cardiometabolic outcomes

#### Table S2. Associations between dietary patterns and cardiometabolic outcomes in proteomics subcohorts

#### Table S3. Associations between dietary patterns and cardiometabolic outcomes in metabolomics subcohorts

#### Table S4. Protein signatures associated with dietary patterns across cohorts

#### Table S5. Summary of replicated protein signatures associated with dietary patterns

#### Table S6. Metabolite signatures associated with dietary patterns across cohorts

#### Table S7. Associations between proteins and cardiometabolic outcomes

#### Table S8. Associations between diet-related proteins and cardiometabolic outcomes

#### Table S9. Associations between metabolites and cardiometabolic outcomes

#### Table S10. Mediation effects of proteins in diet-cardiometabolic disease associations in UK Biobank

#### Table S11. Mediation effects of metabolites in diet-cardiometabolic disease associations

#### Table S12. Bi-directional Mendelian randomization analyses between metabolite and protein Levels

#### Table S13. Scoring criteria for the Alternative Mediterranean Diet (AMED) score

#### Table S14. Scoring criteria for the healthful Plant-based Dietary Index (hPDI) score

#### Table S15. Scoring criteria for the Diet Inflammatory Index (DII) score

#### Table S16. Scoring criteria for the Empirical Dietary Index for Hyperinsulinemia (EDIH) score

#### Table S17. Scoring criteria for the Alternate healty eating index (AHEI) score
